# Power Calculation for Noninferiority Stepped-Wedge Cluster Randomized Trials

**DOI:** 10.64898/2026.10.04.26364705

**Authors:** Justin Guo, Chih-Yuan Hsu, Yu Shyr

**Affiliations:** Montgomery Bell Academy, Nashville, TN 37205, USA; Department of Biostatistics, Vanderbilt University Medical Center, Nashville, TN 37203, USA; Center for Quantitative Sciences, Vanderbilt University Medical Center, Nashville, TN 37203, USA

**Author notes:** Justin Guo and Chih-Yuan Hsu contributed equally to this work. Correspondence: Yu Shyr.

**Keywords:** intraclass correlation, mixed-effects model, noninferiority test, superiority test, stepped-wedge cluster randomized trial

## Abstract

**Background:** Stepped-wedge cluster randomized trials (SW-CRTs) are increasingly used when interventions need to be implemented sequentially across clusters. Despite extensive developments in the design and analysis of superiority SW-CRTs, methods for planning noninferiority SW-CRTs remain limited.

**Methods:** We propose swNIsim, a simulation-based framework for power calculation in noninferiority SW-CRTs using generalized linear mixed-effects models (GLMMs). The proposed approach accommodates both continuous and binary outcomes and supports practical design features, including unequal numbers of clusters across sequences, varying cluster-period sizes, delayed intervention effects, and analyses with or without adjustment for secular time effects. Noninferiority margins can be specified using several commonly applied strategies.

**Results:** Through extensive simulation studies, we examined the impact of key design parameters on statistical power, including the number of periods, the number of clusters per sequence, the number of individuals per cluster-period, and the degree of between-cluster variability. For a fixed total sample size, designs with more time periods consistently achieved higher power. Increasing the number of clusters per sequence produced modest gains in power, whereas greater between-cluster variability reduced power. Type I error rates were well controlled across the scenarios examined. Balanced allocation of clusters across sequences was more efficient than unbalanced allocation schemes. Two real-world SW-CRT examples were used to illustrate application of the method.

**Conclusions:** The proposed swNIsim provides a flexible and practical approach for power calculation in noninferiority SW-CRTs. By accommodating a wide range of design configurations and outcome types, and through implementation as a freely available R package, swNIsim may facilitate the planning and design of noninferiority stepped-wedge trials.

## 1 Introduction

A Stepped-Wedge Cluster Randomized Trial (SW-CRT) is an alternative to the conventional parallel cluster randomized design in which clusters (e.g., hospitals, clinics, or communities) initially receive the control condition and subsequently cross over to the intervention at predefined, staggered time points until all clusters have been exposed. This design is particularly advantageous when simultaneous implementation across all sites is impractical or when withholding the intervention from certain clusters for the entire study would be ethically or operationally challenging [1]. Examples include quality-improvement initiatives, clinic-based screening programs, and workflow modifications that require phased implementation because of limitations in training, staffing, equipment, or administrative capacity [1]. By accommodating gradual rollout, the SW-CRT enables rigorous evaluation of intervention effects under real-world implementation constraints.

A key statistical characteristic of the stepped-wedge design is that each cluster contributes data under both control and intervention conditions. This structure supports both between-cluster and within-cluster comparisons. Because each cluster serves, in part, as its own control, within-cluster comparisons can improve estimation precision, although the magnitude of this gain depends on factors such as the timing of intervention rollout, cluster size, and the underlying correlation structure [2].

Hussey and Hughes [3] established a linear mixed-effects model (LMM) framework for the design and analysis of SW-CRTs. Subsequent work by Hemming et al. [1,4] investigated design variations and their impact on statistical power, while later studies extended these methods to generalized linear mixed-effects models (GLMMs) for non-Gaussian outcomes [5]. Generalized estimating equations have also been proposed as a population-averaged alternative that avoids specifying a distribution for random effects [6].

Despite these methodological advances, existing approaches have been developed for superiority trials, whose objective is to determine whether an intervention performs better than the control. In contrast, methods for noninferiority SW-CRTs remain limited. Noninferiority designs are often appropriate when a new intervention may be slightly less effective than the standard treatment but offers other important advantages, such as reduced cost, easier implementation, improved accessibility, or fewer adverse effects.

To address this methodological gap, we propose swNIsim, a simulation-based framework for power calculation in noninferiority SW-CRTs using GLMMs. Unlike analytical approaches that often rely on restrictive assumptions, simulation-based power calculation provides flexibility for accommodating complex design features commonly encountered in practice. The proposed approach supports both continuous and binary outcomes, allows unequal numbers of clusters across sequences and varying cluster-period sizes, accommodates delayed intervention effects, and permits analyses with or without adjustment for time-period effects. In addition, swNIsim provides flexible options for specifying noninferiority margins. Using extensive simulations, we investigate the effects of major design characteristics on power and Type I error control. We further demonstrate the application of the proposed method through two real-world trial examples and provide an accompanying R package to facilitate its implementation.

## 2 Methods

Suppose the study consists of *T* time periods, corresponding to *T* − 1 intervention sequences. Each sequence s contains *n*_*s*_ clusters, *s* = 1, …, *T* − 1, yielding a total of 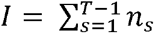 clusters. Every cluster is randomly assigned to one of the *T* − 1 sequences. For clusters assigned to sequence *s*, individuals remain under the control condition through period *s* and cross over to the intervention beginning in period *s* + 1 (Figure 1). Let *K*_*ij*_ denote the number of individuals in cluster *i* during period *j, i* = 1, …, *I* and *j*= 1, …, *T*. For simplicity, we assume a constant cluster size, *K*_*ij*_ =*K* for all *i* and *j*. However, in some settings, such as those involving loss to follow-up or incomplete observations arising from staggered participant enrollment, *K*_*ij*_ may decrease over time.

**Figure 1.**
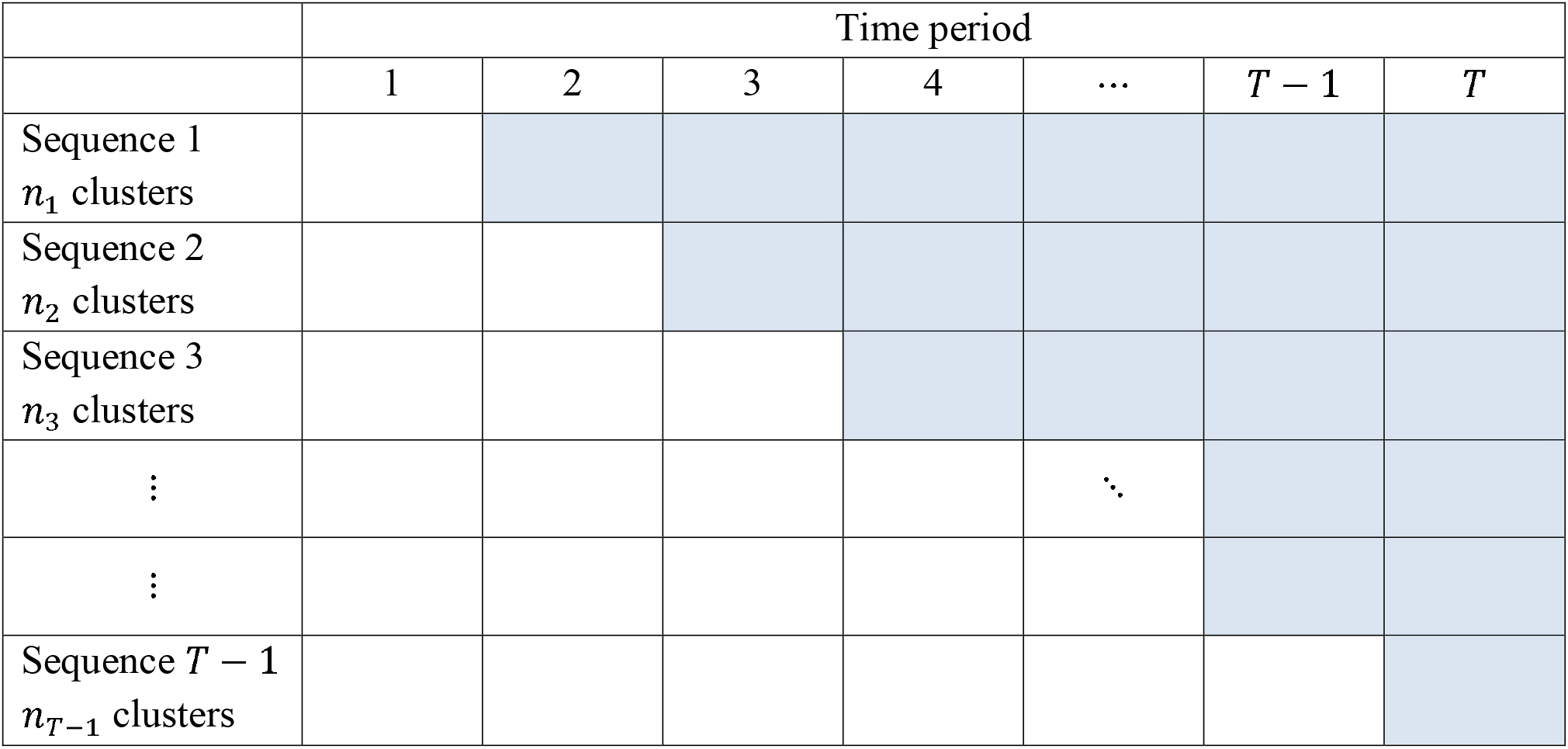
A stepped-wedge design comprising *T* time periods and *T* − 1 sequences, with *n*_*s*_ clusters for each sequence. Clusters in sequence s initiate the intervention at time period *s* + 1.

### 2.1 Statistical models for different outcome types

#### 2.2.1 Continuous outcomes

Let 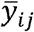 denote the cluster–period mean, defined as the average outcome among individuals in cluster *i* at time period *j*, for *i* = 1, …, *I* and *j*= 1, …, *T*. We assume

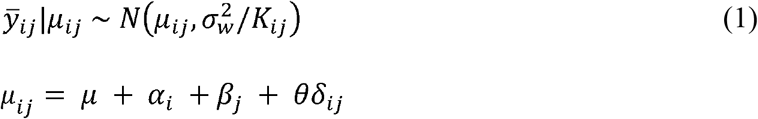

where the cluster-specific random effect *α*_*i*_ follows a normal distribution 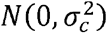. The parameters 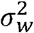 and 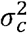 denote the within-individual and between-cluster variances, respectively. The intraclass correlation (ICC), quantifying the correlation between individuals from the same cluster, is given by 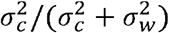. In this model, *µ* denotes the baseline mean under the standard treatment, and *β*_*j*_ is the fixed effect of time period *j*. The parameter *θ* denotes the treatment effect, defined as the difference between the intervention and the standard treatment. The term *δ*_*ij*_ reflects the level of intervention exposure for cluster *i* at time period *j*, allowing for the possibility of delayed intervention effects. In the absence of delayed intervention effects, *δ*_*ij*_ =1 for *j* ≥ *t*_*i*_ and *δ*_*ij*_ =0 otherwise, where *t*_*i*_ denotes the time period at which cluster *i* initiates the intervention. If delayed intervention effects are present, *δ*_*ij*_ may be defined, for example, as *δ*_*ij*_ = (*j* − *t*_*i*_ + 1)/(*T* − 1) for *j* ≥ *t*_*i*_ and *δ*_*ij*_ =0 for *j* < *t*_*i*_.

The noninferiority hypotheses are formulated as

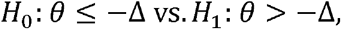

where Δ > 0 denotes the noninferiority margin. A Wald-type test statistic 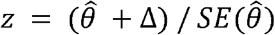 is used to test the one-sided hypotheses, where 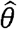 is the estimated treatment effect of *θ*, obtained using a linear mixed-effect model that includes a cluster-level random intercept and, by default, adjusts for fixed effects of time periods, with 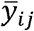 weighted by *K*_*ij*_. The corresponding p-value is 1 − Φ(*z*), where Φ denotes the cumulative distribution function of the standard normal distribution. Noninferiority is concluded when the p-value is less than the prespecified significance level.

Statistical power is estimated using Monte Carlo simulation with a large number of replicates. For each replicate, a dataset is generated under the prespecified design, the planned mixed-effects model is fitted with the R package *lme4* [7], and the noninferiority test is performed. The estimated power is the proportion of replicates in which the null hypothesis is rejected. The swNIsim reports the model-fitting failure rate. By default, replicates with failed model fits are treated as non-rejections and are therefore included in the denominator when calculating power.

#### 2.1.2 Binary outcomes

Let *s*_*ij*_ denote the cluster–period total of binary outcomes, defined as the number of individuals with a response in cluster *i* at time period *j*, for *i* = 1, …, *I* and *j* = 1, …, *T*. We assume

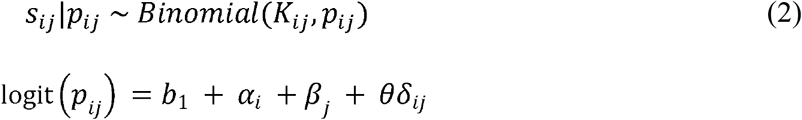

where the cluster-specific random effect *α*_*i*_ follows a normal distribution 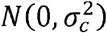. The intercept *b*_1_ is the baseline log odds under the standard treatment, and 1+ exp(−*b*_1_)^−1^ is the corresponding baseline response probability. Time period effects, treatment exposure, and the remaining parameters are defined as in the continuous-outcome model. For this logistic random-intercept model, the latent-scale ICC is given by 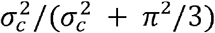 [8]. For example, *σ*_*C*_ = 1 corresponds to a latent-scale ICC of approximately 0.233.

The same one-sided Wald framework is used for the binary outcome. Specifically, 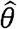 is estimated using a logistic mixed-effects model with a cluster-specific random intercept and, by default, fixed effects for time periods. The noninferiority margin Δ is defined on the log-odds scale. Noninferiority is concluded when 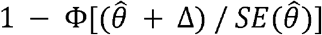 is less than the prespecified significance level. As in the continuous-outcome setting, statistical power is estimated using Monte Carlo simulation with the same rejection and failure-handling rules.

### 2.2 Noninferiority margin determination

A noninferiority margin, Δ on the fitted-model scale is specified through the “margin” argument. Following [9], we provide two options for determining the nonnegative margin Δ.

#### 2.2.1 Continuous outcomes

##### Preserved fraction of the mean under the standard treatment

The aim is for the intervention group to preserve at least a fraction *f*_1_ of the mean response under the standard treatment, where 0 < *f*_1_ < 1. The preserved fraction *f*_1_ represents the proportion of the standard treatment mean that is considered acceptable for the intervention group. This can be expressed as (*μ* + *θ*) > *f*_1_*μ*, which implies *θ* > − (1 − *f*_1_) *μ*. Thus, the margin is Δ = (1 − *f*_1_) *μ*.

##### Preserved fraction of the mean difference between the standard treatment and placebo

The aim is for the intervention to be superior to the placebo and to preserve at least a fraction *f*_2_ of the mean difference between the standard treatment and placebo, where 0 < *f*_2_ < 1. The preserved fraction *f*_2_ represents the proportion of this difference that is considered acceptable for the mean difference between the intervention and placebo. This can be expressed as (*μ* + *θ*) − *μ*_0_ > 0 and (*μ* + *θ*) − *μ*_0_ > *f*_2_(*μ* − *μ*_0_), where *μ*_0_ is the placebo mean. Typically, *μ* − *μ*_0_ > 0 holds, so the margin is Δ = (1 − *f*_2_)(*μ* − *μ*_0_).

### 2.2.2 Binary outcomes

For binary outcomes, the treatment effect is quantified by the log odds ratio. Therefore, the noninferiority margin is specified on the log-odds scale. If a target margin is initially defined on the proportion scale, it will be converted to the corresponding log-odds scale. Throughout this article, a larger proportion indicates greater clinical benefit.

#### Preserved fraction of the success rate under the standard treatment

The aim is for the intervention group to preserve at least a fraction *f*_1_ of the success rate, *p*_1_, of the standard treatment, where 0 < *f*_1_ < 1. The preserved fraction *f*_1_ represents the proportion of the standard treatment success rate that is considered acceptable for the intervention group. The lowest acceptable success probability for the intervention is 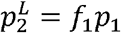. Therefore, the resulting margin on the log-odds scale is 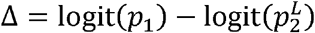.

#### Preserved fraction of the success rate difference between the standard treatment and placebo

The aim is for the intervention to be superior to the placebo and to preserve at least a fraction *f*_2_ of the success rate difference between the standard treatment and placebo, where 0 < *f*_2_ < 1. The preserved fraction *f*_2_ represents the proportion of *p*_1_ − *p*_0_ (standard vs. placebo) that should be retained in *p*_2_ − *p*_0_ (intervention vs. placebo). The lowest acceptable success probability for the intervention is 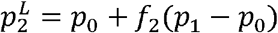. Therefore, the resulting margin on the log-odds scale is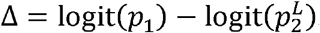.

## 3 Results

### 3.1 Simulation studies

We conducted a series of simulation studies to evaluate the impact of key design characteristics on statistical power, including 1) the number of time periods, 2) the number of clusters per sequence, 3) the number of individuals per cluster-period, and 4) the magnitude of between-cluster variability, while keeping the total number of individuals fixed. Specifically, we considered a range of stepped-wedge design configurations defined by different combinations of the number of time periods (*T*), the number of clusters per sequence (*n*_*s*_ = *n*, for all *s*), and the number of individuals per cluster-period (*K*_*ij*_ = *K*, for all *i, j*): (*T, n, K*) = (7, 10, 20), (7, 4, 50), (6, 12, 20), (6, 8, 30), (5, 15, 20), (5, 10, 30), (4, 20, 20), (4, 10, 40). Each configuration included 1,200 individuals. The significance level was set at 0.025.

The true treatment effect and time period effects were set to zero. i.e., *θ* = 0 and *β*_*j*_ = 0 for all *j*. For Model (1) with continuous outcomes, we set *μ* = 5, *σ*_*w*_ = 2, and *σ*_*C*_ = 1.0, 1.3, or 1.6, corresponding to ICC values of approximately 0.20, 0.30, and 0.39, respectively. The noninferiority margin was set to 0.15. For Model (2) with binary outcomes, we set *b*_1_ = 0.5, corresponding to a conditional response probability of 0.622 under the reference condition, and *σ* _*C*_ = 1.0, 1.5, and 2.0, corresponding to latent-scale ICC values of approximately 0.233, 0.406, and 0.549, respectively. The noninferiority margin was set to 0.18. Empirical power in each scenario was calculated using 100,000 replicates.

For both continuous and binary outcomes, simulations showed that, given the fixed total sample size, designs with a larger number of time periods generally produced higher power (Tables 1 and 2). Increasing the number of clusters per sequence while reducing the number of individuals per cluster-period slightly improved power, whereas greater between-cluster variability reduced power. The number of time periods appeared to be the most influential design factor.

**Table 1.**
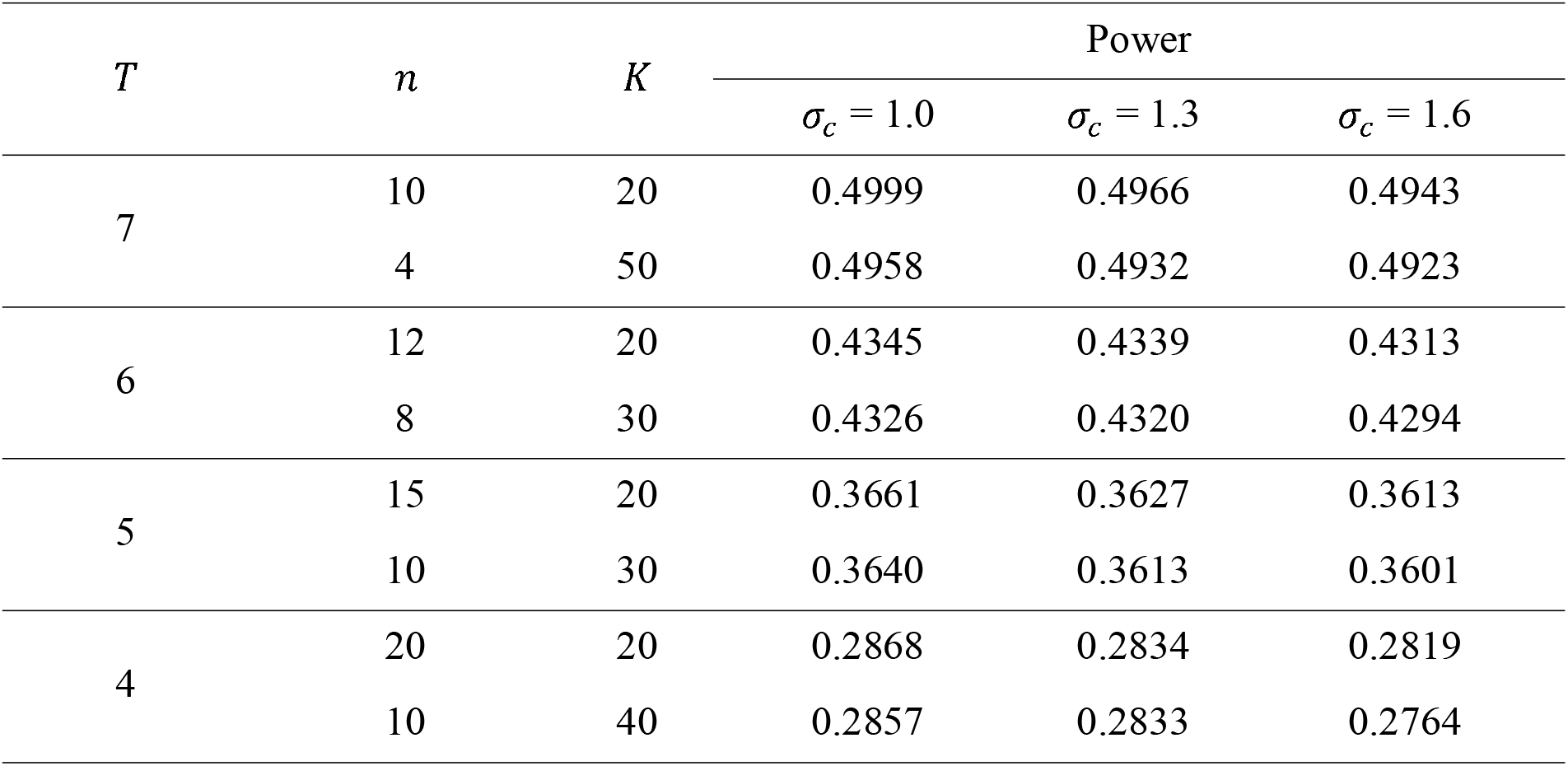
Power for continuous outcomes under models with adjusting for period effects. *μ* = 5, *σ*_*c*_ = (1.0, 1.3, 1.6), *σ*_*w*_ = 2, *θ* = 0, and the noninferiority margin = 0.15. A one-sided *α* = 0.025 was used.

**Table 2.** Power for binary outcomes under models with adjusting for period effects. Baseline log odds *b*_1_ = 0.5, *σ*_*c*_ = (1.0, 1.5, 2.0), *θ* = 0, and the noninferiority margin = 0.18. A one-sided *α* = 0.025 was used.

| $T$ | $n$ | $K$ | Power | | |
| --- | --- | --- | --- | --- | --- |
| | | | $\sigma_c = 1.0$ | $\sigma_c = 1.5$ | $\sigma_c = 2.0$ |
| 7 | 10 | 20 | 0.5570 | 0.4935 | 0.4357 |
|  | 4 | 50 | 0.5493 | 0.4867 | 0.4308 |
| 6 | 12 | 20 | 0.4890 | 0.4295 | 0.3783 |
|  | 8 | 30 | 0.4853 | 0.4259 | 0.3768 |
| 5 | 15 | 20 | 0.4118 | 0.3602 | 0.3161 |
|  | 10 | 30 | 0.4079 | 0.3575 | 0.3137 |
| 4 | 20 | 20 | 0.3213 | 0.2811 | 0.2466 |
|  | 10 | 40 | 0.3158 | 0.2775 | 0.2453 |

We examined Type I error rates under the boundary null hypothesis (*θ* = −Δ). At the nominal one-sided level of 0.025, empirical Type I error rates ranged from 0.0254 to 0.0269 for continuous outcomes and from 0.0250 to 0.0256 for binary outcomes, indicating that the Type I error rate was well controlled (Supplemental Tables 1 and 2). The greatest inflation was observed in the smallest four-period, 30-cluster configuration for continuous outcomes.

We also conducted simulations allowing the number of clusters to vary across sequences. Specifically, for a design with five sequences (*T* = 6), we considered three cluster allocations: (*n*_1_,…, *n*_5_) = (8, 8, 8, 8, 8), (4, 6, 8, 10, 12), and (12, 10, 8, 6, 4). Other parameter settings were identical to those described above, with *K* = 20, and *σ*_*c*_ = 1.3 for continuous outcomes and 1.5 for binary outcomes (Table 3). The highest power was observed under the balanced design, (*n*_1_,…, *n*_5_) = (8, 8, 8, 8, 8), indicating that equal allocation of clusters across sequences is the most efficient configuration.

**Table 3.** Power under scenarios with unequal numbers of clusters across sequences. All models adjusted for period effects. *T* = 6, *K* = 20, *θ* = 0, and (*n*_1_,…, *n*_4_) = (8, 8, 8, 8, 8), (4, 6, 8, 10, 12), or (12, 10, 8, 6, 4), yielding 40 total clusters and 4,800 total observations. For continuous outcomes, *μ*= 5, *σ*_*w*_ = 2, *σ*_*c*_ = 1.3, and the noninferiority margin was 0.15; while for binary outcomes, *b*_1_ = 0.5, *σ*_*c*_ = 1.5, and the noninferiority margin was 0.18. A one-sided *α* = 0.025 was used.

| **Outcome** | **Sequence allocation** | **Clusters by sequence** | **$\sigma_c$** | **Power** |
| --- | --- | --- | --- | --- |
| Continuous | Balanced | 8, 8, 8, 8, 8 | 1.3 | 0.3164 |
| Continuous | Increasing | 4, 6, 8, 10, 12 | 1.3 | 0.2944 |
| Continuous | Decreasing | 12, 10, 8, 6, 4 | 1.3 | 0.2960 |
| Binary | Balanced | 8, 8, 8, 8, 8 | 1.5 | 0.3152 |
| Binary | Increasing | 4, 6, 8, 10, 12 | 1.5 | 0.2872 |
| Binary | Decreasing | 12, 10, 8, 6, 4 | 1.5 | 0.2854 |

Because no secular time effects were imposed (*β*_*j*_ = 0 for all *j*), we also evaluated power when period effects were not included in Models (1) and (2). As shown in Tables 4 and 5, the overall power patterns were similar to those obtained from models that adjusted for period effects. However, power was consistently higher when period effects were excluded, reflecting the efficiency loss incurred by estimating time-effect parameters that were truly zero.

**Table 4.** Power for continuous outcomes under models without adjusting for period effects. μ = 5, *σ*_*c*_ = (1.0, 1.3, 1.6), *σ*_*w*_ = 2, *θ* = 0, and the noninferiority margin = 0.15. A one-sided *α* = 0.025 was used.

| $T$ | $n$ | $K$ | Power | | |
| --- | --- | --- | --- | --- | --- |
| | | | $\sigma_c = 1.0$ | $\sigma_c = 1.3$ | $\sigma_c = 1.6$ |
| 7 | 10 | 20 | 0.8529 | 0.8522 | 0.8519 |
|  | 4 | 50 | 0.8508 | 0.8513 | 0.8504 |
| 6 | 12 | 20 | 0.8048 | 0.8032 | 0.8027 |
|  | 8 | 30 | 0.8034 | 0.8020 | 0.8015 |
| 5 | 15 | 20 | 0.7417 | 0.7408 | 0.7391 |
|  | 10 | 30 | 0.7402 | 0.7406 | 0.7387 |
| 4 | 20 | 20 | 0.6643 | 0.6631 | 0.6617 |
|  | 10 | 40 | 0.6627 | 0.6604 | 0.6607 |

**Table 5.** Power for binary outcomes under models without adjusting for period effects. Baseline log odds *b*_1_ = 0.5, *σ*_*c*_ = (1.0, 1.5, 2.0), *θ* = 0, and the noninferiority margin = 0.18. A one-sided *α* = 0.025 was used.

| $T$ | $n$ | $K$ | Power | | |
| --- | --- | --- | --- | --- | --- |
| | | | $\sigma_c = 1.0$ | $\sigma_c = 1.5$ | $\sigma_c = 2.0$ |
| 7 | 10 | 20 | 0.8996 | 0.8515 | 0.7964 |
|  | 4 | 50 | 0.8955 | 0.8476 | 0.7910 |
| 6 | 12 | 20 | 0.8596 | 0.8034 | 0.7421 |
|  | 8 | 30 | 0.8570 | 0.8007 | 0.7391 |
| 5 | 15 | 20 | 0.8021 | 0.7424 | 0.6760 |
|  | 10 | 30 | 0.8011 | 0.7380 | 0.6743 |
| 4 | 20 | 20 | 0.7292 | 0.6636 | 0.5991 |
|  | 10 | 40 | 0.7231 | 0.6594 | 0.5938 |

### 3.2 Real Applications

Two real-world stepped-wedge cluster-randomized trials were used to illustrate the proposed swNIsim. These examples serve as parameterized demonstrations of the methodology rather than exact reproductions of the original studies or re-evaluations of their superiority-based sample size calculations. The first example is based on the TheraPPP trial for continuous-outcome setting [10]; the second example is based on the PRIM-ER trial for binary-outcome setting [11,12].

#### 3.2.1 Continuous outcome example: TheraPPP trial

The TheraPPP trial evaluated the effectiveness of a standardized care pathway for mechanically ventilated (MV) patients, including subgroups with hypoxemic respiratory failure (HRF) and acute respiratory distress syndrome (ARDS). The primary continuous outcome was 28-day ventilator-free days (VFDs). This trial was conducted across 17 adult intensive care units (ICU) in Alberta, Canada. In the full stepped-wedge design, 16 ICUs were organized into eight groups, each containing two ICUs, with one cluster initiating the intervention every two months.

For the overall MV patients, the study protocol reported a baseline mean of 21 VFDs, a standard deviation of 10 days, an ICC of 0.15. The original trial was designed for 90% power at a two-sided alpha of 0.05 to detect a 0.9-day superiority difference [10]. For illustration of the proposed noninferiority framework, we adopted these design parameters and specified *T* − 1 = 8 (sequences), *n*_*s*_ = 2 (number of clusters per sequence), *μ* = 21, *σ*_*w*_ = 10, *σ*_*c*_ = 4.2, *β*_*j*_ = 0, and *θ* = 0. The noninferiority margin was defined as Δ = (1 − *f*_1_) = 4.2, where *f*_1_ = 0.8. Under these assumptions, the required cluster-period size *K*_*ij*_ = *K* = 5 was determined to achieve 90% power at a one-sided significance level of alpha = 0.025. Similarly, for the subgroup of patients with sustained ARDS, the protocol reported a baseline mean of 11 VFDs, a standard deviation of 10 days, an ICC of 0.01. Using these parameters for methodological illustration, we specified *T* − 1 = 8, *n*_*s*_ = 2, = 11, *σ*_*w*_ = 10, *σ*_*c*_ = 1, *β*_*j*_ = 0, and *θ* = 0. The noninferiority margin was defined as Δ = (1 − *f*_1_) = 2.2, where *f*_1_ = 0.8. The required cluster-period size *K*_*ij*_ = *K* = 14 was determined to achieve 90% power at a one-sided significance level of alpha = 0.025.

#### 3.2.2. Binary outcome example: PRIM-ER trial

The PRIM-ER trial evaluated the effect of a multicomponent intervention to initiate palliative care in the emergency department (ED) on hospital admission. The primary outcome was hospital admission following an ED visit. Based on the assumptions used in the original sample size calculation (a baseline hospital admission rate of 70%, an ICC of 0.10, and 30 participating ED sites), the trial was estimated to have approximately 85% power to detect an absolute difference of 3 percentage points in hospital admission rates.

For illustration of the proposed noninferiority framework, we adopted these design parameters and specified *T* − 1 = 30, *n*_*s*_ = 1, *b*_1_ = − 0.847, corresponding to a baseline non-admission probability of *p*_1_ = 0.3, *σ*_*c*_ = 0.605, determined to yield a latent-scale ICC of 0.1, and *β*_*j*_ = 0, and e = 0. The noninferiority margin was defined as Δ = 0.305, derived using *f*_1_ = 0.8 and *p*_1_ = 0.3. Under these assumptions, the required cluster-period size *K*_*ij*_ = *K* = 6 was determined to achieve 85% power at a one-sided significance level of alpha = 0.025.

## 4 Discussion

In this study, we proposed swNIsim, a simulation-based framework for power calculation in noninferiority stepped-wedge cluster randomized trials. While extensive methodological research has focused on the design of superiority SW-CRTs, methods for planning noninferiority SW-CRTs remain limited. The proposed framework addresses this gap by providing a flexible approach for estimating power under generalized linear mixed-effects models for both continuous and binary outcomes. The method accommodates several practical design features frequently encountered in applied studies, including unequal numbers of clusters across sequences, varying cluster-period sizes, delayed intervention effects, and analytic models with or without adjustment for period effects.

The simulation studies provide several insights into the design of noninferiority SW-CRTs. For a fixed total sample size, increasing the number of time periods consistently improved power for both continuous and binary outcomes. This finding suggests that the number of periods is a particularly influential design parameter in stepped-wedge trials. Increasing the number of clusters per sequence while reducing the number of individuals per cluster-period resulted in modest improvements in power, whereas increasing between-cluster variability reduced power.

We also found that balanced allocation of clusters across sequences provided higher power than unequal allocation schemes, supporting the use of balanced designs whenever feasible.

The simulations further demonstrated adequate control of the Type I error rate across the scenarios examined. Although only slight inflation was observed, the largest deviations occurred in designs with relatively few clusters. This observation is consistent with previous studies reporting that asymptotic inference in mixed-effects models may exhibit inflated Type I error rates when the number of clusters is small [13,14]. Consequently, investigators should exercise caution when designing SW-CRTs with a limited number of clusters and may wish to evaluate operating characteristics through simulation before finalizing the design.

Another notable finding concerns adjustment for secular time effects. When no true period effects were present in the data-generating mechanism, analyses that omitted period effects achieved higher power than models that included them. This result reflects the loss of efficiency associated with estimating unnecessary parameters. In practice, however, secular trends often exist and should generally be incorporated into the primary analysis. Therefore, the observed gains in power should not be interpreted as a recommendation to routinely omit period effects, but rather as an illustration of the trade-off between model complexity and efficiency.

Several limitations should be noted. The proposed approach models correlations among observations from the same cluster across periods through a shared cluster-level random intercept. Although this assumption is widely used in the stepped-wedge literature, it may not adequately capture more complex correlation structures, such as decay or autoregressive patterns over time [15,16], that arise in some applications. Future extensions could incorporate alternative correlation structures and additional random effects [17,18]. In addition, simulation-based methods are computationally more intensive than analytical approaches, particularly when large numbers of replicates are required to obtain precise power estimates. Nevertheless, simulation-based approaches offer greater flexibility and can accommodate complex design features and modeling assumptions that are difficult to address analytically.

## 5 Conclusions

We propose swNIsim, a simulation-based framework for power calculation in noninferiority stepped-wedge cluster randomized trials. The proposed method enables investigators to evaluate realistic design scenarios and estimate power under a wide range of assumptions that may be difficult to address analytically. Implemented as a freely available R package, swNIsim provides a practical and flexible tool for the planning and design of noninferiority SW-CRTs.

## Data Availability

Additional file 1: Online Supplementary Material. A freely available R package swNIsim is at https://github.com/justing-uo/swNIsim.

## Abbreviations

ARDS: acute respiratory distress syndrome
ED: emergency department
GLMMs: generalized linear mixed-effects models
HRF: hypoxemic respiratory failure
ICC: intraclass correlation
ICU: intensive care units
LMM: linear mixed-effects model
MV: mechanically ventilated
SW-CRT: stepped-wedge cluster randomized trial
VFDs: ventilator-free days

## Declarations

### Ethics approval and consent to participate

Not applicable.

### Consent for publication

Not applicable.

### Availability of data and materials

Additional file 1: Online Supplementary Material. A freely available R package *swNIsim* is at https://github.com/justing-uo/swNIsim.

### Authors’ contributions

JG and CYH were major contributors in writing the manuscript and developing the R package. YS was the major contributor in the conception and design of the work.

